# A low pre-existing anti-NS1 humoral immunity to DENV is associated with microcephaly development after gestational ZIKV exposure

**DOI:** 10.1101/2024.05.07.24306978

**Authors:** Sebastián Castro-Trujillo, William R. Mejía, Katherine Segura, Rocío Vega, Doris Salgado, Carlos E. Fonseca, Ángela M. Ortiz, Federico Perdomo-Celis, Irene Bosch, Carlos F. Narváez

## Abstract

**Background:** Gestational Zika virus (ZIKV) infection is associated with the development of congenital Zika syndrome (CZS), which includes microcephaly and fetal demise. The magnitude and quality of orthoflavivirus-specific humoral immunity have been previously linked to the development of CZS. However, the role of ZIKV NS1-specific humoral immunity in mothers and children with prenatal ZIKV exposure and CZS remains undefined. In addition, considering that most of the at-risk population lives in dengue virus (DENV)-endemic areas, it is not clear what is the association between pre-existing DENV NS1-specific humoral immunity and CZS.

**Methods:** Here, we studied 328 mothers and children with a clinical diagnosis and seropositivity for ZIKV infection during pregnancy, included during the 2015-2016 ZIKV epidemic in Colombia. We also performed clinical evaluation and pediatric neurological follow-up. The relative levels of circulating NS1-specific IgM and IgG against ZIKV and DENV were evaluated in mothers and children, and the association with the development of microcephaly was analyzed.

**Results:** DENV and ZIKV IgG-NS1 antibodies in pregnant women were placentally transferred, and this passage and its duration in children depended on the maternal levels of the antibodies. We reported that higher concentrations of pre-existing DENV, but not ZIKV IgG-NS1 antibodies, were associated with a reduced risk of CZS-related microcephaly. Also, we observed that the IgM-NS1 response presents as long-term and has a minor association with poor outcomes.

**Conclusions:** The development of microcephaly in children prenatally exposed to ZIKV is associated with low plasma levels of placentally transferred, pre-existing DENV IgG-NS1 antibodies. These data are compatible with a protective role of anti-NS1 IgG antibodies against ZIKV infection during pregnancy and highlight the promising role of NS1 as an orthoflavivirus vaccine target in high-risk populations.

## INTRODUCTION

Zika Virus (ZIKV) is an arbovirus transmitted to humans through the bite of *Aedes* mosquitoes [1]. Like other orthoflaviviruses (such as dengue virus [DENV]), ZIKV is an enveloped virus with a positive-sense, single-stranded RNA genome that codifies for three structural proteins (E, preM, and C) and seven nonstructural proteins [2]. Zika virus is a re-emerging pathogen that in 2022 had affected at least 89 countries worldwide, primarily in tropical and subtropical regions [3]. Between October 2015 and July 2016, active autochthonous transmission of ZIKV was confirmed in Colombia, leading to a nationwide epidemic declaration [4]. In February 2016, the World Health Organization (WHO) declared ZIKV infection a public health emergency of international concern [3], given the link between ZIKV infection during pregnancy and subsequent birth defects [5]. Indeed, although ZIKV infection is asymptomatic in most cases or courses as a self-limited and mild disease [6], when infection occurs during pregnancy, ZIKV can be vertically transmitted, resulting in spontaneous abortion or the development of congenital Zika syndrome (CZS) [6]. This neonatal syndrome includes microcephaly, intracranial calcifications, ophthalmologic abnormalities, and neurodevelopmental disorders [7]. In addition, infants with CZS may have a case fatality rate of 10% within the first years of life [8]. In Colombia, microcephaly prevalence increased from 2.1 per 10,000 live births in 2015 to 9.6 in 2016, supporting the association between reported ZIKV infections and the occurrence of this clinical entity [9].

The mechanisms responsible for the development of CZS are not fully understood. While it is known that ZIKV can infect and induce the death of human neural progenitor cells [10,11], tissue damage may also be associated with an exacerbated immune response triggered by the infection [12–14]. Furthermore, the humoral immune response against ZIKV and pre-existing immunity to orthoflaviviruses may play protective or pathogenic roles in CZS [15]. Indeed, previous studies have shown that neutralizing human antibodies directed to the ZIKV E protein can protect against ZIKV replication and fetal demise in mice [16]. In contrast, pre-existing DENV-specific antibodies in mice infected with ZIKV during pregnancy may enhance vertical ZIKV transmission, resulting in microcephaly [17]. It is worth noting that while there has been extensive research on the humoral response to the ZIKV and DENV E proteins, the response to the NS1 protein has not been studied as extensively. This protein is highly immunogenic [18] and is associated with severe disease as well as placental dysfunction [19]. A considerable proportion of the antibodies elicited by ZIKV and DENV are targeted toward NS1 [20–22]. These antibodies exhibit lower cross-reactivity than those directed to the E protein of related orthoflaviviruses, particularly DENV, which makes them a valuable tool in the diagnosis [23]. In addition, anti-ZIKV NS1 antibodies decrease the risk of antibody-dependent enhancement of infection [20,24], highlighting their role in antibody-mediated protection against disease and as an optimal target in vaccine development. In keeping with these data, previous studies have suggested that anti-ZIKV NS1 antibodies derived from adult samples have a protective role against ZIKV disease [24,25]. Moreover, it was demonstrated that antibodies recognizing cell-surface NS1 confer protection against ZIKV in pregnant mice [26]. However, it is unknown the role of anti-DENV NS1 antibodies in CZS.

Previous research has examined the link between the cellular immune response in mothers and children and the development of CZS [27,28]. However, the connection between pre-existing humoral immunity in mothers and CZS in their children remains unexplored. Particularly, it is unknown whether anti-ZIKV NS1 humoral immunity in mothers and children with prenatal ZIKV exposure is associated with CZS, especially in severe forms such as microcephaly. In addition, since most of the at-risk population lives in dengue-endemic areas, it is essential to determine the association between pre-existing anti-DENV NS1 antibodies and children and CZS development. To investigate these aspects, we evaluated the levels of anti-NS1 IgM and IgG antibodies against ZIKV and DENV in mothers and their offspring exposed to ZIKV during pregnancy. We observed the transplacental passage of anti-NS1 IgG (but not IgM) antibodies from ZIKV and DENV, with an expected clearance rate. In addition, we found a relationship between the levels of pre-existing anti-NS1 humoral immunity to DENV and the occurrence of CZS-related microcephaly.

## METHODS

### Ethics statement

The study adhered to the principles of the Declaration of Helsinki and received approval from the Ethics, Bioethics, and Research Committee of the Hospital Universitario Hernando Moncaleano Perdomo in Neiva, Huila, Colombia (No. 004-007, April 19, 2016) and from the Ethics and Research Committee of clínica ESIMED (Minute No. 007 dated September 14, 2016). All participating mothers provided written informed consent for themselves and their children.

### Participants, samples, and pediatric follow-up

A multicenter study was conducted including two independent cohorts with a total of 436 participants: the Hospital Universitario Hernando Moncaleano Perdomo cohort (n=272; mothers and children), and the clínica ESIMED cohort (n=164; mothers and children), both located in Neiva, southern Colombia, a hyper-endemic region for dengue [29]. We included mother/child binomials with the antecedent of gestational exposure to ZIKV during the 2015-2016 Zika epidemic. Exposure to ZIKV was defined as the presence of plasma anti-ZIKV NS1 IgG antibodies in the mothers, accompanied by signs and symptoms of Zika disease during pregnancy at the peak of the epidemic period. In all cases, TORCH syndrome (Toxoplasmosis, Other infections [such as syphilis and HIV], Rubella, Cytomegalovirus, Herpes simplex) was ruled out by serological or molecular testing. The study inclusion time was 0.2 to 19 months after the child’s birth. At enrollment, a blood sample was obtained simultaneously from all mothers and their infants (4 or 2 mL, respectively, collected in ethylenediaminetetraacetic acid-containing tubes). Plasma was isolated through centrifugation at 200 *x g* for 5 minutes and stored at -80°C until analysis. Neurological, speech therapy, and ophthalmological assessments were conducted for 113 children up to 12 months after enrollment. The childreńs cognitive, language, and motor development were evaluated with the Bayley-III child development scale and applied by a pediatric neurologist specialist. Neurodevelopment alteration was defined as <85 points in any of the parameters evaluated [30]. The complete characterization of neurological disturbances in these children can be found elsewhere [31]. The study chart flow is presented in **Figure 1**.

**Figure 1.**
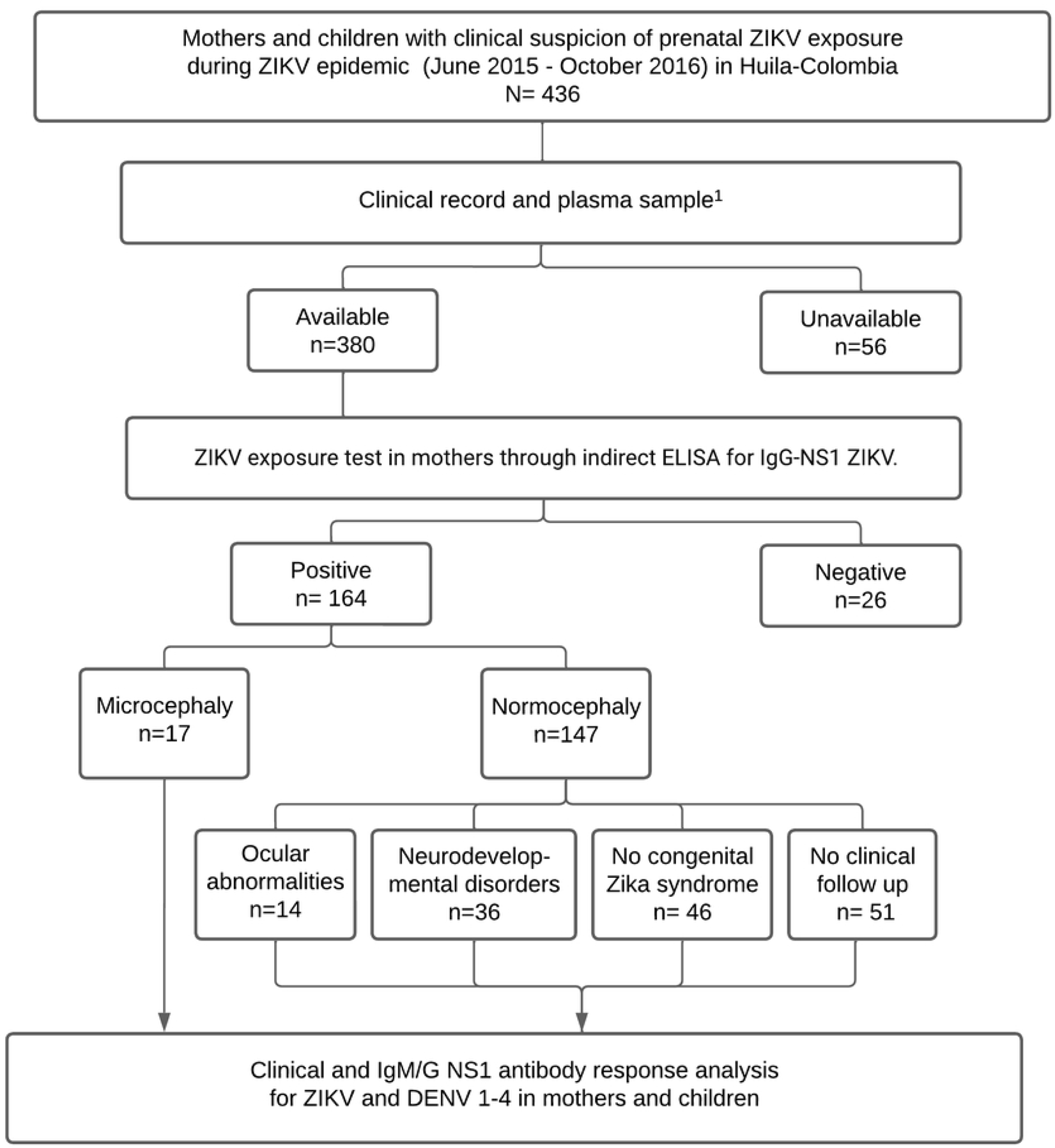
Study flowchart. ^1^The plasma sample was taken after birth in the clinical follow-up.

### Measurement of plasma anti-NS1 IgM and IgG antibodies to ZIKV and DENV

Indirect enzyme-linked immunosorbent assays (ELISA) based on recombinant whole NS1 protein of ZIKV (Surinam strain, No. ZIKV-NS1) and a mix of the four NS1 DENV serotypes (DENV-1 Nauru/Western Pacific/1974, DENV-2 Thailand/16681/84, DENV-3 Sri Lanka D3/H/IMTSSA/SRI/2000/1266 and DENV-4 Dominica/814669/1981) were used to evaluate the relative levels of IgM and IgG antibodies in plasma samples (here referred to as IgG-NS1 or IgM-NS1), as previously reported [32,33]. Samples obtained before 2014 from healthy individuals serologically without exposure to either ZIKV or DENV were used as a negative control. Based on the negative control, a detection limit of 0.27 and 0.36 optical densities (OD 450 nm) was established for anti-NS1 IgM and IgG, respectively. Additionally, samples from convalescent patients with confirmed ZIKV or DENV infection were included as a positive control. Anti-NS1 IgM and IgG antibodies are reported as a positivity rate (percent of samples with OD 450 nm above the detection limit) and relative levels (raw OD 450 nm values). All samples were evaluated in duplicate. Inter-plate variability in the positive and negative controls was lower than 15%.

### Statistical Analysis

Statistical analysis was performed with GraphPad Prism version 10. Quantitative analyses are presented as medians and range. Frequency analyses were done using Fisher’s exact test. The Mann-Whitney and Wilcoxon tests were used for unpaired and paired analyses of the two groups, respectively. Three or more independent groups were compared with Kruskal-Wallis and Dunn’s post-hoc tests. The correlation between variables was determined using the Spearman rank test. The cutoff of DENV1-4/ZIKV IgG-NS1 ratio to predict microcephaly outcome was evaluated using receiver operating characteristic (ROC) curve analyses. The better cutoff was chosen using the Youden J Index (sensitivity + specificity -1). A p-value <0.05 was considered significant. When comparing IgM or IgG values, statistical analysis was performed only if the median level of any group was above the limit of detection of the assay.

## RESULTS

### Characteristics of the study cohort

We included 218 mother and child pairs with clinical suspicion of prenatal ZIKV exposure. From them, 190 had available clinical records and plasma samples. In turn, 164 mothers were positive for IgG-NS1 ZIKV (**Figure 1**). Clinical follow-up was performed in 113 children. From them, 67 cases presented CZS: 17 developed microcephaly, 14 had ocular abnormalities, and 36 exhibited only neurodevelopmental impairment (**Figure 1**). In 51 normocephalic infants, clinical follow-up could not be performed. The clinical and demographic characteristics of the study cohort are shown in **Table 1**. We grouped the participants as those without or with microcephaly. We did not observe differences in the age of mothers from both groups (**Table 1**). However, consistent with the link between ZIKV infection during the first trimester of pregnancy and microcephaly [34], in our cohort, a higher proportion of cases of microcephaly occurred after ZIKV infection in the first trimester (**Table 1**). In contrast, most cases of neurodevelopment and ocular alterations, such as optical nerve hypoplasia and chorioretinitis, occurred after ZIKV infection during the second trimester (**Supplementary Table 1**). We did not observe differences in the gestational age at birth, in the clinical manifestations of suspected ZIKV infection in the mothers (including fever, exanthema, and myalgias/arthralgias), and in the biological sex of the infants between those with versus without microcephaly (**Table 1**).

### Placental transfer of ZIKV and DENV IgG-NS1 antibodies

The dynamic of the humoral response to ongoing gestational ZIKV infection, as well as pre-existing antibodies against DENV, remains unclear. Thus, we evaluated the plasma levels of ZIKV and DENV IgM-NS1 and IgG antibodies in pregnant mothers and their newborns to determine the transfer of these antibodies across the placenta and their temporal kinetics. Consistent with the absence of ongoing or recent ZIKV or DENV infection at the time of sample collection, overall, there were low or undetectable levels of IgM-NS1 both in mothers and children (**Figure 2A** and B; **Supplementary Figure 1A**). This was also reflected in a poor correlation between IgM-NS1 in mothers and children (**Figure 2A** and **B**). Moreover, consistent with the lack of transplacental passage of IgM [35], in some mothers with detectable levels of DENV IgM-NS1, these values were not observed in their children (**Figure 2B**).

**Figure 2.**
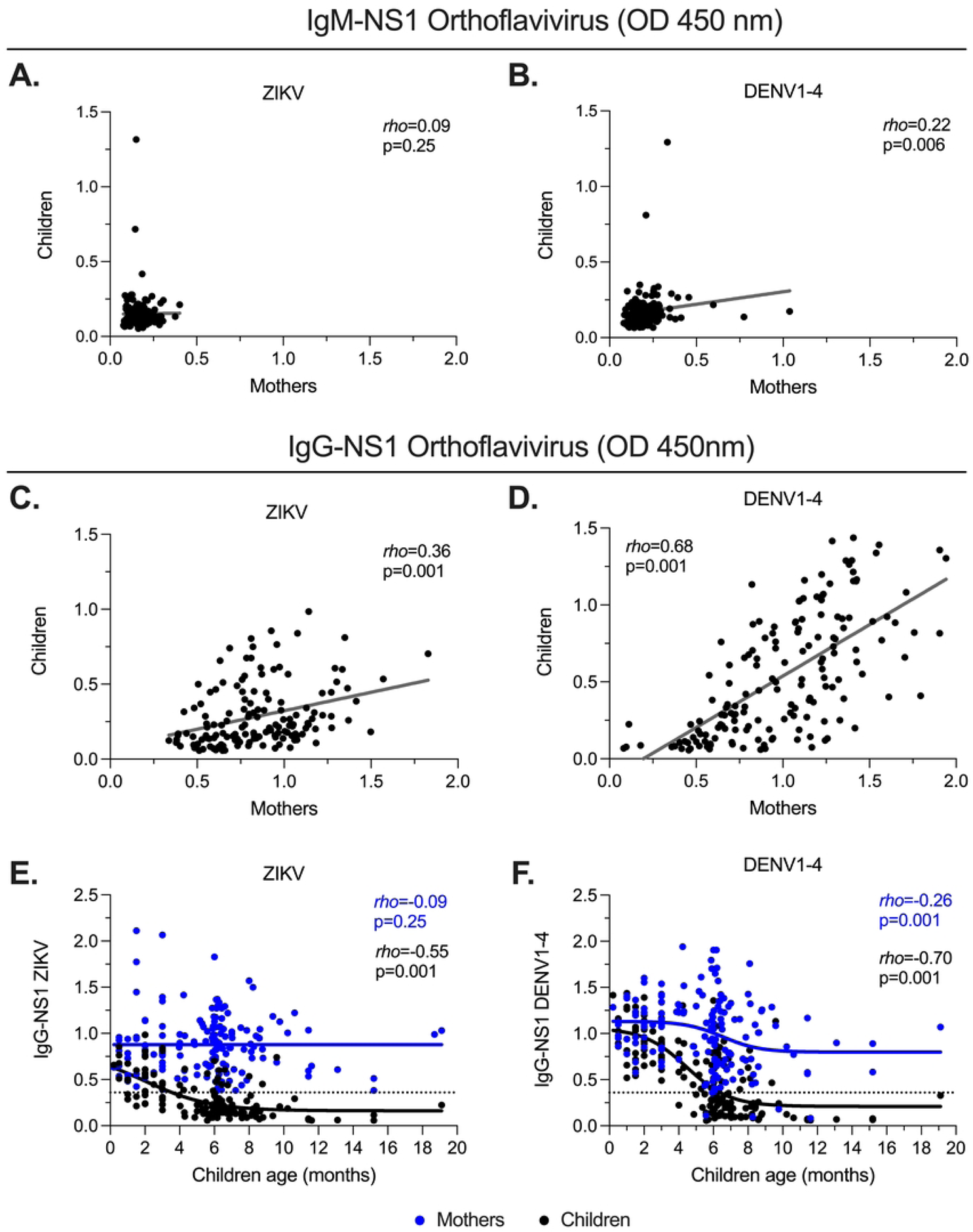
Placentally-transferred IgG-NS1. **A-B.** Correlation analysis of the relative plasma levels of ZIKV (**A**) and DENV1-4 (**B**) IgM-NS1 between mothers and children. **C-D.** Correlation analysis of the relative levels of ZIKV (**C**) and DENV1-4 (**D**) IgG-NS1 between mothers and children. **E-F.** Correlation and non-linear regression analysis of the relative levels of ZIKV (**E**) and DENV1-4 (**F**) IgG-NS1 in relation to the children age. In all the cases, the *rho* and p-value of the Spearman test are shown. The dashed lines indicate the ELISA limit of detection.

In contrast to IgM-NS1, there was a high positivity rate of IgG-NS1, a pattern consistent with the high levels of endemic DENV transmission in this population [29]. As expected, the levels were higher in the mothers than in the children (**Supplementary Figure 1B**). In addition, we observed higher levels of DENV than ZIKV IgG-NS1 in mothers and children (**Supplementary Figure 1B**). It is known that placental transport of IgG depends on maternal levels [35]. In keeping with this notion, we observed a positive correlation between IgG-NS1 in mothers and children, but the strength of association was higher for DENV IgG-NS1 (**Figure 2C** and **D**). Consistent with the kinetics of clearance of maternally acquired anti-orthoflavivirus IgG [36,37], the levels of ZIKV and DENV IgG-NS1 declined in the infants predominantly from the sixth month of age onward, nearly disappearing by the eighth month (**Figure 2E** and **F**). Of note, in children before six months of age, the frequency of undetectable IgG-NS1 was significantly higher for ZIKV than for DENV (69% vs. 30%, p<0.001), indicating that a significant proportion of infants could be susceptible to ZIKV reinfection early after birth. In contrast, the levels of these antibodies remained relatively stable in the mothers (**Figure 2E** and **F**).

We also evaluated the impact of the time of ZIKV exposure on the levels of DENV and ZIKV IgG-NS1. As such, we compared the antibody levels in samples from mothers and children exposed to ZIKV during the first versus the third trimester of pregnancy. The mothers who were exposed to ZIKV in the third trimester had significantly higher levels of DENV, but not ZIKV, IgG-NS1 than those exposed in the first trimester (**Supplementary Figure 2A** and B), reflecting the decay of DENV-specific antibody levels in the latter. A similar trend was observed for the children (**Supplementary Figure 2C and D**). These data also indicate different dynamics and/or sources of DENV- and ZIKV-specific antibodies. Indeed, we did not observe a correlation between ZIKV and DENV IgG-NS1 antibody levels in plasma from mothers (*rho*=-0.05; p=0.45) or children (*rho*=0.004; p=0.95). These results are likely related to the modest effect of prior DENV infections on the breadth and magnitude of ZIKV-reactive memory B cell responses [38]. In summary, these findings confirm the passive transfer of maternal anti-orthoflavivirus IgG-NS1 (but not IgM-NS1) antibodies and their subsequent clearance around the sixth month after birth.

### Poor association between ZIKV and DENV IgM-NS1 antibodies and CZS-related microcephaly

We next evaluated the relationship between ZIKV and DENV IgM-NS1 antibodies and the development of microcephaly in children with CZS. Consistent with the low positivity rate in most of the cases (**Figure 2A** and **B; Supplementary Figure 1A**), we did not observe significant differences in ZIKV or DENV IgM-NS1 in the mothers of children with microcephaly versus those without microcephaly, both in terms of the positivity rate (**Figure 3A**), or the relative plasma levels (**Figure 3B**). In contrast, we observed that children with microcephaly exhibited a higher seropositivity rate for ZIKV IgM-NS1 than those without microcephaly (**Figure 3C**). This suggests a long-lasting IgM response in children with microcephaly, considering the early prenatal exposure to ZIKV. However, the plasma levels of these antibodies (as well as those against DENV) in general were below the detection limit (**Figure 3D**). Thus, overall, there is a minor association between the IgM-NS1 response and the development of CZS-related microcephaly, and the ZIKV IgM-NS1 response is long-lasting in a fraction of the children.

**Figure 3.**
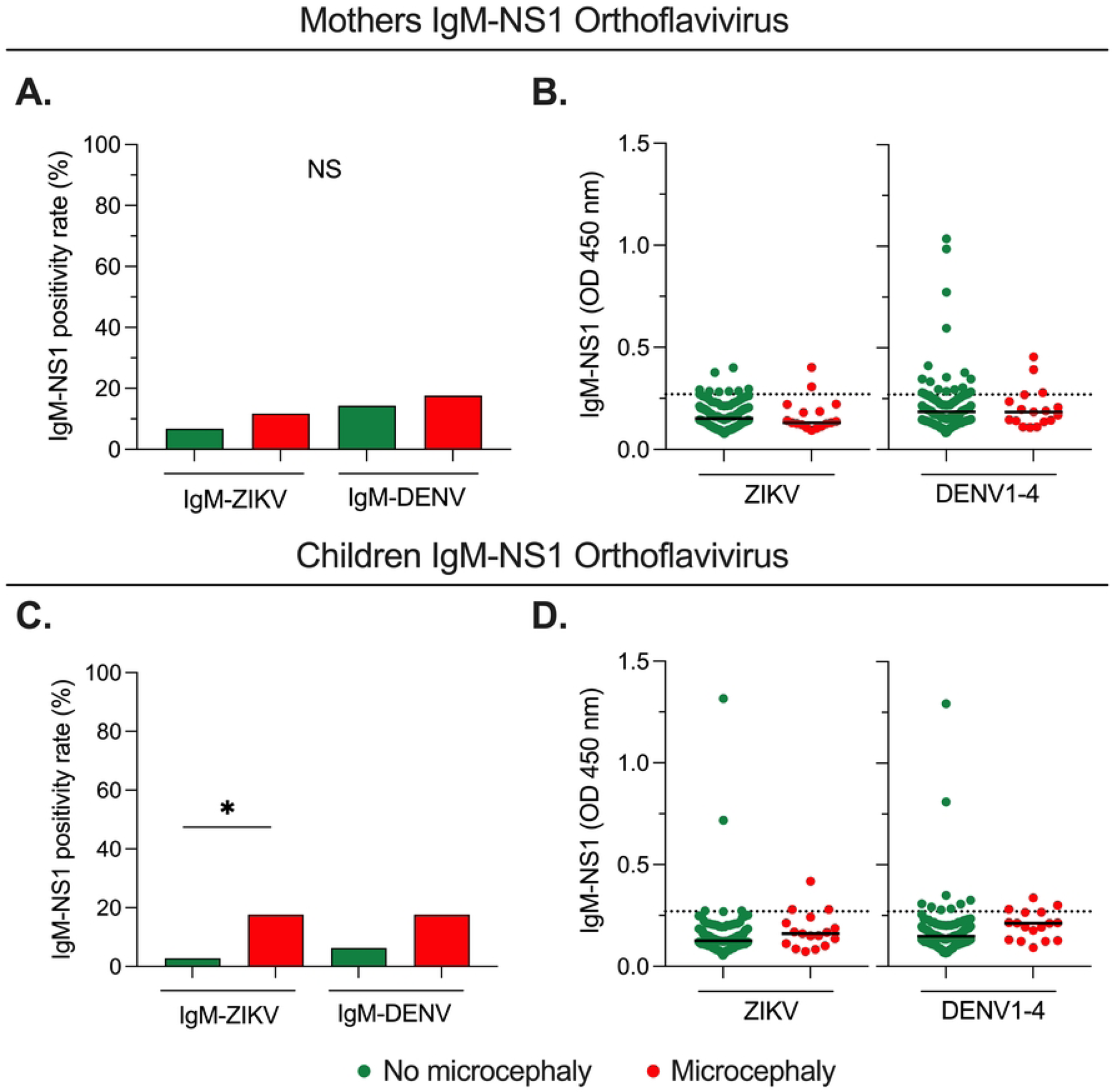
Poor association between IgM-NS1 antibodies and CZS-related microcephaly. **A.** and **C.** Frequency of ZIKV and DENV1-4 IgM-NS1 positivity rate in mothers (**A**) and children (**C**) with or without microcephaly; Fisher’s test. **B** and **D.** Relative levels of ZIKV and DENV1-4 IgM-NS1 in mothers (**B**) and children (**D**) with or without microcephaly; Mann-Whitney test. The dashed lines indicate the ELISA limit of detection. NS: Not statistically significant. *p<0.05.

### Low pre-existing DENV IgG-NS1 is associated with CZS-related microcephaly

We examined the association between orthoflavivirus IgG-NS1 antibody levels in mothers and children and the development of microcephaly. We observed that mothers of children with microcephaly had a lower positivity rate for DENV IgG-NS1 in comparison with those without microcephaly (**Figure 4A**). Of note, this comparison was not performed for ZIKV IgG-NS1 since positivity for this test was a study inclusion criterion. When we evaluated the relative levels of IgG-NS1, we observed significantly lower levels of DENV, but not ZIKV, antibodies in mothers of children with microcephaly relative to those without microcephaly (**Figure 4B**). In addition, we employed the DENV/ZIKV IgG-NS1 ratio to measure the predominant viral-specific humoral response. This analysis indicated that mothers of children with microcephaly had a lower DENV/ZIKV IgG-NS1 ratio in comparison with those without microcephaly (**Figure 4C**). Interestingly, a ROC curve analysis indicated that the DENV/ZIKV IgG-NS1 ratio in samples from the mothers is robust in discriminating between both groups of children (**Figure 4D**).

**Figure 4.**
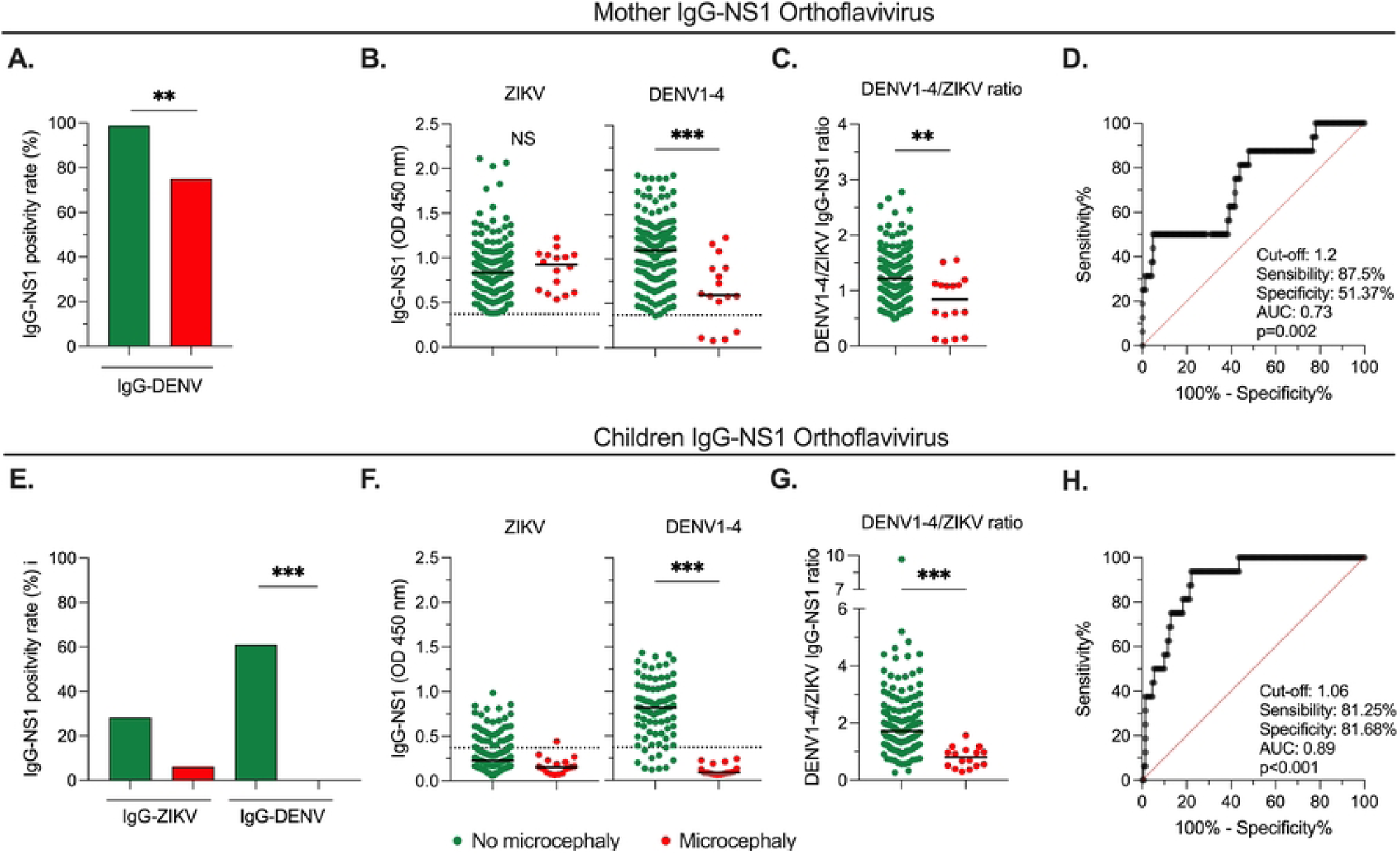
Association between IgG-NS1 antibodies and CZS-related microcephaly. **A.** Frequency of DENV1-4 IgG-NS1 positivity rate in mothers of children with or without microcephaly. **B.** Relative plasma levels of ZIKV and DENV1-4 IgG-NS1 in mothers of children with or without microcephaly. **C-D.** DENV1-4/ZIKV IgG-NS1 ratio in mothers of children with or without microcephaly (**C**), and the respective ROC curve analysis (**D**). **E.** Frequency of ZIKV and DENV1-4 IgG-NS1 positivity rate in children with or without microcephaly. **F.** Relative plasma levels of ZIKV and DENV1-4 IgG-NS1 in children with or without microcephaly. **G-H.** DENV1-4/ZIKV IgG-NS1 ratio in children with or without microcephaly (**G**), and the respective ROC curve analysis (**H**). In A and E, the p-value of the Fisher’s test is shown. In B, C, F, and G, the p-value of the Mann-Whitney test is shown. The dashed lines indicate the ELISA limit of detection. NS: Not statistically significant. **p<0.01; ***p<0.001.

We also evaluated the association between IgG-NS1 in plasma from the children exposed prenatally to ZIKV and the risk of microcephaly. Of note, the samples from children who developed microcephaly were obtained at an older age (median [range] of 8.2 months [4-15]) than those without microcephaly (median [range]: 6 months [1-19]; P=0.001). Thus, considering the clearance of maternally-transferred antibodies beyond 6 months of age (**Figures 2E** and **F**) and that passively transferred antibodies depended on maternal levels (**Figure 2C** and **D**), it was expected observe that children with microcephaly had a lower positivity rate for DENV IgG-NS1 (**Figure 4E**), as well as lower relative levels of DENV-specific antibodies (**Figure 4F**), in comparison with those without microcephaly. No differences were obtained for ZIKV IgG-NS1 antibodies (**Figure 4E** and **F**). The DENV/ZIKV IgG ratio analysis abrogates the bias introduced by the difference in the age of children with and without microcephaly. We observed a significantly lower DENV/ZIKV IgG-NS1 ratio in children with microcephaly in comparison with non-microcephalic children (**Figure 4G**), and this ratio could discriminate between both groups of children (**Figure 4H**).

Finally, we evaluated the levels of DENV and ZIKV IgM/IgG-NS1 in mothers and children without microcephaly but who developed neurodevelopmental or ocular alterations and compared them with those who did not develop CZS. Overall, we did not detect DENV and ZIKV IgM-NS1 in all the groups analyzed (**Supplementary Figure 3A and C**). Moreover, when we evaluated IgG-NS1, contrary to what we observed for microcephaly cases, we obtained comparable levels of these antibodies in mothers of children with neurodevelopmental or ocular alterations relative to those without CZS (**Supplementary Figure 3B**). Mothers from children with neurodevelopmental alterations had even significantly higher DENV IgG-NS1 antibodies than those from the no CZS group (**Supplementary Figure 3B**). A similar non-significant trend was obtained in the children (**Supplementary Figure 3D**).

In summary, low levels of pre-existing DENV IgG-NS1 antibodies are associated with the development of microcephaly, but not neurodevelopmental or ocular alterations, in children with CZS.

## DISCUSSION

Developing safe and effective immunization strategies for women of reproductive age living in orthoflavivirus endemic areas is a public health priority given the risks associated with the vertical transmission of ZIKV [15]. In addition, identifying immune correlates of protection against CZS is highly needed. Here, we observed that pre-existing DENV IgG-NS1 antibodies in pregnant women are placentally transferred to their children, and this passage depends on the maternal levels of the antibodies. In addition, in the setting of ZIKV infection during pregnancy, higher levels of these antibodies are associated with a reduced risk of CZS-related microcephaly but not with the occurrence of neurodevelopmental or ocular alterations. In contrast, a minor association was observed for the IgM-NS1 response, which is most likely explained by the absence of transplacental passage of this isotype [35]. Consistent with these data, a previous study that prospectively followed a cohort of 1,453 adults in dengue-endemic areas demonstrated that a two-fold increase in IgG titers against DENV NS1 is associated with a 9% reduction in the risk of ZIKV infection and symptoms [39]. Thus, pre-existing immunity directed to the NS1 protein of orthoflaviviruses seems to play a protective role against ZIKV infection and CZS-related microcephaly.

Due to its absence in viral particles, NS1 is usually not considered a vaccine target to induce humoral responses against orthoflaviviruses. However, secreted and cell-associated forms of NS1 can be found, and both have varying immunogenic properties [18]. For instance, around 30% of ZIKV- and DENV-reactive antibodies bind to NS1 [20,22]. Moreover, multiple studies have shown a protective role of anti-NS1 against orthoflavivirus infection. As such, seminal work showed that immunization with DENV-2 NS1 confers protection against a homologous DENV infection in mice [40]. More recent studies obtained similar findings in mice immunized with a ZIKV NS1-based vaccine. Specifically, this vaccine elicited high titers of protective antibodies, and passive serum transfer from vaccinated mice conferred protection against lethal challenge [41]. In addition, antibodies directed to the cell-surface form of ZIKV NS1 may confer protection against ZIKV infection during pregnancy by a mechanism related to complement deposition and opsonization [26]. Similarly, monoclonal antibodies directed to the cell-surface form of West Nile Virus NS1 activate Fcγ receptor-mediated phagocytosis and clearance of infected cells [42], while anti-ZIKV NS1 antibodies promote antibody-dependent cellular toxicity [43,44]. On the other hand, ZIKV NS1 induces placental dysfunction, increasing vascular permeability and spreading the virus. Thus, anti-NS1 antibodies could also protect against ZIKV infection by decreasing the immune evasion effects and placental dysfunction induced by the secreted form of NS1 [19,45].

The previous studies indicate a protective role of anti-NS1 homotypic antibodies. However, it is unclear the role of anti-NS1 cross-reactive antibodies in individuals with pre-existing immunity to DENV and subsequent ZIKV infection. In this regard, it is worth mentioning that the NS1 proteins from DENV and ZIKV share some degree of homology [46,47], and some monoclonal antibodies against DENV NS1 have cross-reactivity to ZIKV NS1 [48,49]. Importantly, these anti-DENV NS1 cross-reactive antibodies can mediate protective effects against ZIKV NS1-induced endothelial dysfunction [48], as well as decrease viral replication and cytokine production *in vitro* [49]. In contrast, other studies have shown that the depletion of DENV-reactive antibodies in serum does not significantly affect ZIKV neutralization [38,50], suggesting that type-specific antibodies are responsible for most of this antibody-mediated protection. Considering the lack of antibody-dependent enhancement of ZIKV infection by anti-NS1 antibodies [48], these studies support the notion that cross-reactive antibodies targeting DENV NS1 may provide protection against ZIKV infection and are a helpful vaccine target to prevent CZS.

In addition to the direct effects of anti-NS1 antibodies, they could be a surrogate of other immune mechanisms of protection against CZS. For instance, ZIKV NS1-based immunization in immunocompetent mice was recently shown to elicit high levels of anti-ZIKV NS1 antibodies. However, T cell–mediated immunity, rather than anti-NS1 antibodies, is crucial for protecting against the ZIKV challenge in this model [51]. Alternatively, efficient cooperation between anti-DENV NS1 antibodies and CD4^+^ T cells may be required for optimal protection upon NS1-based vaccination [52]. Moreover, the presence of antibodies to anti-ZIKV NS1 antibodies correlates with the presence of ZIKV neutralizing antibodies, the latter probably being the direct protective mechanism against infection [25]. In summary, anti-NS1 antibodies are an important arm of humoral immunity against orthoflaviviruses. Their effector mechanisms and biological relevance of homotypic and cross-reactive anti-NS1 antibodies in the setting of ZIKV infection and CZS remain to be determined.

Several studies have documented neurodevelopment and ocular alterations in infants born to mothers exposed to ZIKV during pregnancy, even in the absence of microcephaly [53,54]. However, the mechanisms behind this clinical entity in the setting of ZIKV infection are unknown [55]. Here, we show that these neurological alterations differ respective to microcephaly in CZS both in the time of ZIKV exposure and the association with placentally-transferred humoral immunity. Likely, gestational ZIKV infection in the first trimester directly alters human brain cell development [11], and low levels of protective anti-orthoflavivirus antibodies may predispose to microcephaly. However, ZIKV infection beyond this time (when macroscopic brain architecture has been already developed), along with maternal immune activation upon infection (which could be indicated by the levels of anti-NS1 antibodies), may lead to changes in functional brain connectivity and neurodevelopment alterations [56]. Novel studies should address the role of maternal immune activation and orthoflavivirus humoral immunity in this setting.

Our study has some limitations. We did not prospectively measure antibody responses in pregnant women early after ZIKV exposure, which could be better correlated with clinical outcomes in newborns. However, the results of our study with a high number of mothers/children binomials exposed to prenatal ZIKV infection support the utility of measuring orthoflavivirus humoral immunity to identify individuals with increased risk of severe CZS in endemic areas. Furthermore, it is important to note that children with microcephaly were older than those without this condition at the time of sample analysis. This has implications for the levels of anti-NS1 antibodies. However, the analysis of the DENV/ZIKV IgG-NS1 ratio (which is independent of the age of the child) is useful to control this variable between both groups of individuals. Finally, a limitation of our study is that the mechanisms behind the association found for IgG-NS1 antibodies and microcephaly development were not explored. In this regard, at least two unanswered questions remain to be addressed: which mechanisms are related to the association between anti-DENV NS1 antibodies and a good clinical outcome in CZS? Is the anti-NS1 IgG response a reflect of homologous or heterologous neutralizing antibodies or virus-specific T cells playing a direct role in protection against ZIKV infection?

In conclusion, here we show that the development of microcephaly in children with CZS is associated with low plasma levels of placentally transferred, pre-existing DENV IgG-NS1 antibodies. These data align with the notion that the anti-orthoflavivirus NS1 IgG response may play a protective role against ZIKV infection during pregnancy. These results have important implications for the design of novel orthoflavivirus vaccine candidates and the identification of correlates of protection. As such, an optimal anti-orthoflavivirus NS1 IgG response may be required for adequate protection against prenatal ZIKV infection.

## Data Availability

The figures of the manuscript present all relevant data. The corresponding author will provide additional information upon request.

## Acknowledgments

We thank the children and parents who kindly participated in this study and the Pediatric Department of Hospital Universitario de Neiva, particularly Elisabeth Días. We also thank the “Asociación Milagros de Dios” and Piedad Perilla for technical support.

## Funding information

This study was supported by Universidad Surcolombiana, Vicerrectoría de Investigación grant code 3660, and the MIT International Science and Technology Initiatives (MISTI)-Colombia.

## Notes

### Competing Interest Statement

The authors have declared no competing interest.

### Funding Statement

Yes

